# Detection of Left Ventricular Systolic Dysfunction from Single-Lead Electrocardiography Adapted for Wearable Devices

**DOI:** 10.1101/2022.12.03.22283065

**Authors:** Akshay Khunte, Veer Sangha, Evangelos K Oikonomou, Lovedeep S Dhingra, Arya Aminorroaya, Bobak J Mortazavi, Andreas Coppi, Harlan M Krumholz, Rohan Khera

## Abstract

Artificial intelligence (AI) can detect left ventricular systolic dysfunction (LVSD) from electrocardiograms (ECGs). Wearable devices could allow for broad AI-based screening but frequently obtain noisy ECGs. We report a novel strategy that automates the detection of hidden cardiovascular diseases, such as LVSD, adapted for noisy single-lead ECGs obtained on wearable and portable devices. Overall, 385,601 ECGs were used for development of a standard and noise-adapted model. For the noise-adapted model, ECGs were augmented during training with random gaussian noise within four distinct frequency ranges, each emulating real-world noise sources. Both models performed comparably on clean ECGs with an AUROC of 0.90. The noise-adapted model performed significantly better on the same test set augmented with four distinct real-world noise recordings at multiple signal-to-noise ratios (SNRs), including noise isolated from a portable device ECG. The standard and noise-adapted models had an AUROC of 0.72 and 0.87, respectively when evaluated on ECGs augmented with portable ECG device noise at an SNR of 0.5. This approach represents a novel strategy for the development of wearable adapted tools from clinical ECG repositories.

## INTRODUCTION

Artificial intelligence (AI) can detect left ventricular systolic dysfunction (LVSD) from electrocardiograms (ECGs), a diagnosis that has traditionally relied on comprehensive echocardiography or other cardiac imaging.^1,2^ Even though AI-ECG is a promising screening tool for detecting LVSD, the algorithms have been designed in clinically obtained 12-lead ECGs. Advances in wearable and handheld technologies now enable the point-of-care acquisition of single-lead ECG signals, paving the path for efficient and scalable AI screening tools for use with these technologies.^3,4^ This improved accessibility could enable broader AI-based screening for LVSD, but the reliability of such tools is limited by the presence of noise in data collected from wearable devices.^5^ Consequently, the performance of wearable-based models for detecting LVSD may degrade in the real-world setting, with lower performance than observed in the original single-lead derivatives of the clinical development studies.^6,7^

In the absence of large, labelled datasets of wearable ECGs, the development of algorithms that can detect underlying structural heart disease on wearable devices relies on single-lead information specifically adapted from 12-lead ECGs extracted from clinical ECG libraries. However, this process does not specifically account for the unique data acquisition challenges encountered with wearable ECG, possibly contributing to their inconsistent diagnostic performance. Indeed, several sources of noise exist in wearable data, arising from factors such as poor electrode contact with the skin, movement and muscle contraction during the ECG, and external electrical interference.^8–11^ This noise has practical implications, as models demonstrate poorer performance when tested on all available wearable ECG data as opposed to selected high-quality subsets.^7^ This marked difference in performance based on noise has limited wearable device-based screening programs, with a wearable device-based atrial fibrillation screening study disqualifying 22% of patients due to insufficient signal quality.^5^ Accounting for this noise is a prerequisite to develop broadly accessible models that will form the basis of effective screening programs for LVSD in the community.

In the present study, we hypothesized that a novel, noise-enhanced training approach can boost the performance of wearable-adapted, single-lead ECG models for accurate and noise-agnostic identification of LVSD. Our method, which relies on training on single-lead ECG data derived from clinical ECGs and augmented with custom noise patterns developed in key frequency ranges relevant for specific ECG noise signatures, explicitly accounts for, and generalizes to real-world noise patterns observed in wearable devices.

## RESULTS

### Study Population

There were 2,135,846 consecutive 12-lead ECGs performed at the Yale-New Haven Hospital (YNHH) between 2015 and 2021, 440,072 of which had accompanying TTEs acquired within 15 days of the ECG. We developed the models on 385,601 of the ECG-TTE pairs, representing 116,210 unique patients, who had a complete 12-lead ECG recording (**Figure S1**). The signal from Lead I, the standard lead obtained from wearable devices,^4^ was then isolated from each 12-lead ECG. All selected single-lead ECG recordings contained 10 seconds of Lead I signal at 500 Hz. The single-lead ECGs were then split at the patient level into training, validation, and test datasets (85%-5%-10%).

Of these ECGs, 56,894 (14.8%) were from patients with LVSD, defined as having a paired TTE recording of LV ejection fraction (LVEF) below 40%. Additionally, 40,240 (10.4%) had an LVEF between 40% and 50%, and the remaining 288,467 (74.8%) had an LVEF of 50% or higher. Patients had a median age of 68 years (IQR 56, 78) at the time of ECG recording and 50,776 (43.7%) of the patients were women. A total of 75,928 (65.3%) patients were non-Hispanic white individuals, 14,000 (12.0%) were non-Hispanic Black, 9,349 (8.0%) were Hispanic, and 16,843 (14.5%) were from other racial backgrounds (**Table S1**).

### Detection of LV Systolic Dysfunction

The noise-adapted model was trained on a noise-augmented development set. High and low pass filtering was used to isolate five-minute samples of random gaussian noise within four different frequency ranges encompassing real-world ECG noises, including 3-12 Hz, 12-50 Hz, 50-100 Hz, and 100-150 Hz. The first of these four ranges, 3-12 Hz, was selected to emulate motion artifact noises due to tremors,^12,13^ while the 12-50 Hz and 100-150 Hz frequency ranges encompass more frequently occurring lower- and higher-frequency muscle activation artifacts, respectively.^9,13^ The 50-100 Hz domain was selected to represent electrode motion noises.^13^ Both this domain and the 100-150 Hz frequency range, which contain multiples of 50 and 60 Hz, the two mains frequencies used in ECG acquisition,^9^ also serve to emulate powerline interference noise.^9,13^ These noise samples were then used to generate the noise-augmented development set, in which ECGs were selectively noised with random, 10-second sequences of one of the four frequency-banded gaussian noises at one of four signal-to-noise ratios (SNRs) each time an ECG was loaded. The standard model was trained on the original training set (described in Methods, Isolation of Frequency-Banded Gaussian Noise and Methods, Noise Augmentation).

Both models were trained to detect LVEF below 40%, a threshold present in most heart failure diagnosis guidelines,^14^ and consistent with prior work.^1,15^ With an AUROC for detection of LVEF < 40% of 0.90 (95% CI 0.89-0.91) and 0.90 (95% CI 0.88-0.91), the noise-adapted and standard models, respectively, performed similarly on a held-out test set without added noise (p-value = 0.60). AUPRC on this clean held-out test set was 0.46 and 0.48, respectively. The noise-adapted model had specificity and sensitivity of 0.68 and 0.92, respectively and a PPV and NPV of 0.20 and 0.99, respectively. The standard model had sensitivity and specificity of 0.69 and 0.91, respectively and a PPV and NPV of 0.21 and 0.99, respectively. The noise-adapted model’s performance was comparable to the standard model across subgroups of age, sex, and race (**Table 1**).

**Table 1.** Performance of noise-adapted model on test ECGs without noise across demographic subgroups. Abbreviations: PPV, positive predictive value; NPV, negative predictive value; AUROC, area under receiver operating characteristic curve; CI, confidence interval, AUPRC, area under precision recall curve; OR, odds ratio.

| Labels | PPV | NPV | Specificity | Sensitivity | AUROC (95% CI) | AUPRC | OR |
| --- | --- | --- | --- | --- | --- | --- | --- |
| **All** | 0.203 | 0.990 | 0.676 | 0.924 | 0.896 (0.886-0.905) | 0.455 | 25.195 |
| **Male** | 0.231 | 0.986 | 0.653 | 0.916 | 0.881 (0.867-0.894) | 0.478 | 20.508 |
| **Female** | 0.163 | 0.994 | 0.716 | 0.923 | 0.913 (0.898-0.927) | 0.404 | 30.318 |
| **White** | 0.204 | 0.990 | 0.675 | 0.922 | 0.894 (0.883-0.906) | 0.445 | 24.544 |
| **Black** | 0.211 | 0.990 | 0.596 | 0.945 | 0.880 (0.854-0.906) | 0.461 | 25.407 |
| **Hispanic** | 0.208 | 0.992 | 0.740 | 0.923 | 0.917 (0.881-0.953) | 0.535 | 34.158 |
| **Other** | 0.218 | 0.995 | 0.772 | 0.941 | 0.932 (0.900-0.964) | 0.545 | 54.122 |
| **65 or Older** | 0.198 | 0.989 | 0.606 | 0.936 | 0.879 (0.865-0.893) | 0.432 | 22.611 |
| **Under 65** | 0.209 | 0.990 | 0.731 | 0.910 | 0.908 (0.895-0.921) | 0.481 | 27.543 |

### Standard and Noise-Adapted Model Performance on Noised ECGs

The performance of each model was also tested on four distinct real-world noise recordings, including three half-hour recordings containing electrode motion, muscle artifact, and baseline wander noise sourced from the publicly available MIT-BIH noise stress test database.^9^ Both models were tested on separate versions of the held-out test set augmented with each of these 3 real-world noises at eight different signal-to-noise ratios (described in Methods, Acquisition of Real-World, Public ECG Noise Recordings, and Methods, Noise Augmentation).

For the noise-adapted model, model performance was comparable across all SNRs for each MIT-BIH noise with AUROC between 0.86-0.89, 0.87-0.89, and 0.88-0.89 for electrode motion, muscle artifact, and baseline wander noise, respectively. The standard model had lower performance across all SNRs for every noise, with AUROC between 0.79-0.86, 0.81-0.86, and 0.80-0.86 for electrode motion, muscle artifact, and baseline wander noise, respectively (**Table 2, Figure 1**).

**Table 2.** Performance of noise-adapted and standard model on test ECGs with across different types of noise. Abbreviations: SNR, signal-to-noise ratio, PPV, positive predictive value; NPV, negative predictive value; AUROC, area under receiver operating characteristic curve; CI, confidence interval, AUPRC, area under precision recall curve.

| Model | Noise | SNR | PPV | NPV | Specificity | Sensitivity | AUROC (95% CI) | AUPRC |
| --- | --- | --- | --- | --- | --- | --- | --- | --- |
| *Noise-Adapted* | Clean | N/A | 0.203 | 0.990 | 0.676 | 0.924 | 0.896 (0.886-0.905) | 0.455 |
|  | Portable ECG | 0.5 | 0.152 | 0.993 | 0.523 | 0.956 | 0.871 (0.861-0.882) | 0.392 |
|  | Portable ECG | 2 | 0.188 | 0.992 | 0.636 | 0.940 | 0.889 (0.880-0.899) | 0.439 |
|  | Electrode Motion | 0.5 | 0.126 | 0.993 | 0.395 | 0.971 | 0.858 (0.846-0.869) | 0.361 |
|  | Electrode Motion | 2 | 0.167 | 0.991 | 0.579 | 0.945 | 0.885 (0.875-0.895) | 0.426 |
|  | Muscle Artifact | 0.5 | 0.175 | 0.989 | 0.608 | 0.927 | 0.871 (0.861-0.882) | 0.389 |
|  | Muscle Artifact | 2 | 0.194 | 0.990 | 0.654 | 0.930 | 0.891 (0.881-0.900) | 0.449 |
|  | Baseline Wander | 0.5 | 0.186 | 0.990 | 0.634 | 0.932 | 0.883 (0.873-0.893) | 0.427 |
|  | Baseline Wander | 2 | 0.201 | 0.991 | 0.669 | 0.929 | 0.892 (0.883-0.902) | 0.457 |
| *Standard* | Clean | N/A | 0.207 | 0.988 | 0.688 | 0.910 | 0.895 (0.884-0.905) | 0.475 |
|  | Portable ECG | 0.5 | 0.091 | 0.981 | 0.132 | 0.972 | 0.723 (0.706-0.739) | 0.200 |
|  | Portable ECG | 2 | 0.125 | 0.991 | 0.398 | 0.958 | 0.834 (0.822-0.847) | 0.329 |
|  | Electrode Motion | 0.5 | 0.088 | 0.988 | 0.079 | 0.990 | 0.792 (0.779-0.806) | 0.239 |
|  | Electrode Motion | 2 | 0.152 | 0.988 | 0.537 | 0.925 | 0.855 (0.843-0.866) | 0.333 |
|  | Muscle Artifact | 0.5 | 0.100 | 0.991 | 0.210 | 0.979 | 0.807 (0.795-0.820) | 0.245 |
|  | Muscle Artifact | 2 | 0.181 | 0.988 | 0.631 | 0.912 | 0.864 (0.853-0.875) | 0.344 |
|  | Baseline Wander | 0.5 | 0.095 | 0.993 | 0.158 | 0.987 | 0.802 (0.789-0.816) | 0.236 |
|  | Baseline Wander | 2 | 0.169 | 0.986 | 0.599 | 0.908 | 0.855 (0.843-0.866) | 0.322 |

**Figure 1.**
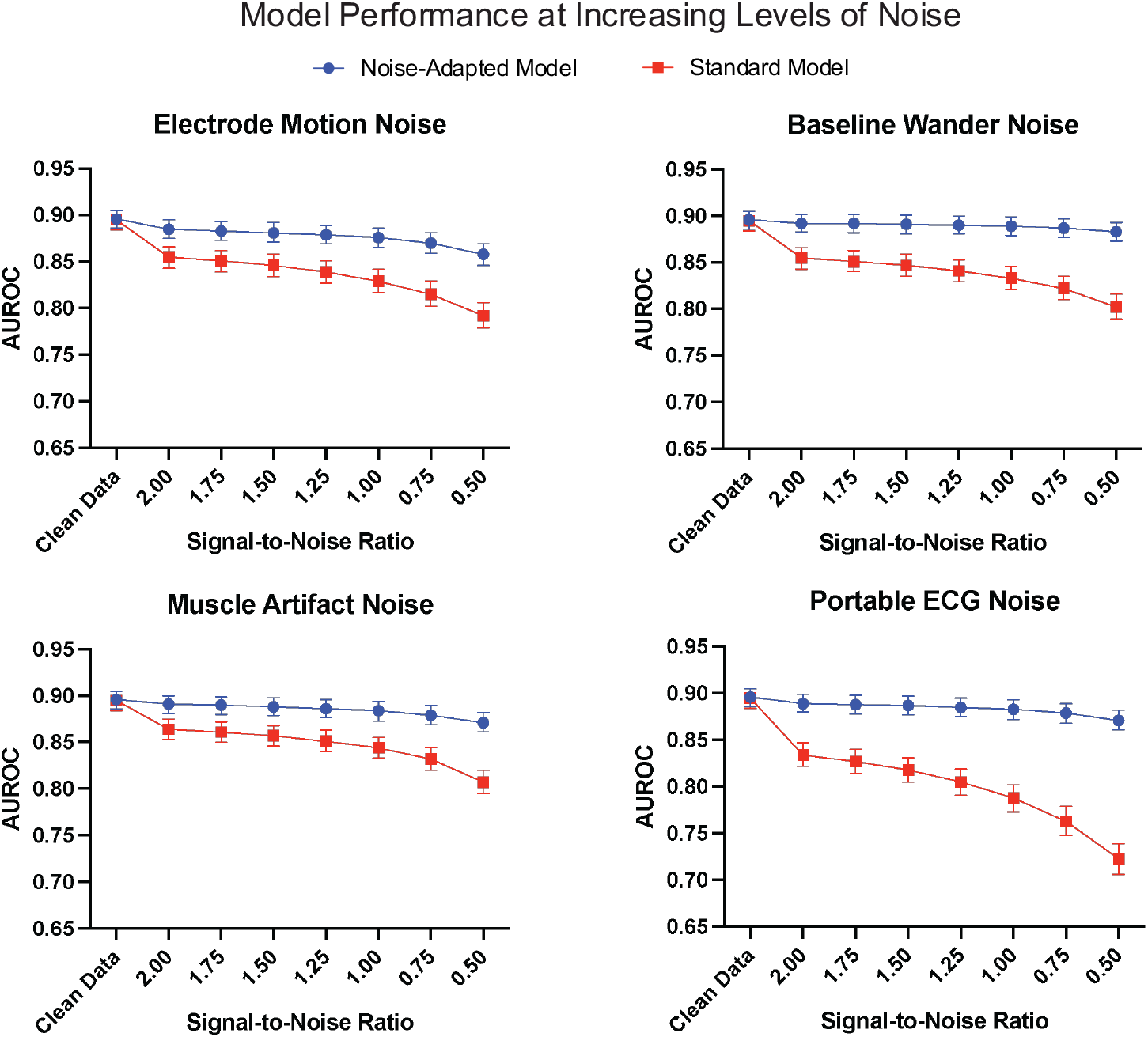
Standard and noise-adapted model performance at increasing levels of noise. Abbreviations: AUROC, area under receiver operating characteristic curve; ECG, electrocardiogram.

### Standard and Noise-Adapted Performance with Portable ECG Device Noise

Real-world portable ECG device noise, isolated using a Fast Fourier Transform-based technique on a 30-second KardiaMobile 6L ECG recording, was also used to evaluate both models (Methods, Noise Extraction from a Portable Device ECG). Each model was used to generate predictions for noise-augmented ECGs across all SNRs, with the noise-adapted model’s AUROC ranging from 0.87 to 0.89. The standard model’s performance was significantly lower at each SNR, ranging from 0.72 to 0.83. This difference was most pronounced at an SNR of 0.5, at which the noise-adapted model retained an AUROC of 0.87 (95% CI 0.86-0.88) and the standard model had an AUROC of 0.72 (95% CI 0.71-0.74, p-value < 0.001) (**Table 2, Figure 1**).

### Explaining Performance Differences in Standard and Noise-Adapted Models

To gain mechanistic insights into the variation in performance for clean and noise-augmented data by different models, we visually and quantitatively assessed differences in the embedding outputs of both noised and clean ECGs for both standard and noise-adapted models. For this, we focused on the output of the 320-dimensional last fully connected layer of each model before generating final predictions. These predictions were collected for five different versions of the same 1,000 ECGs—once with the original ECG, and once for each of the four real-world noises. The variation in these predictions due to the addition of noise was visualized by using uniform manifold approximation and projection (UMAP),^16^ which constructs a two-dimensional representation of the 320-dimension prediction vectors for each noise and model combination. They were also quantitatively assessed using scaled Euclidean distances between prediction vectors for the same ECG with and without each type of noise for both models.

For the standard model, the predictions for each of the noised ECGs were visually distinct from those of the clean ECGs, despite being for the same set of 1,000 ECGs, and differing only on the added noise. However, for the noise-adapted model, there wasn’t a visual separation in the model’s predictions between clean and noised ECGs, indicating that the predictions of the noise-adapted model are more resilient than those of the standard model (**Figure 2**).

**Figure 2.**
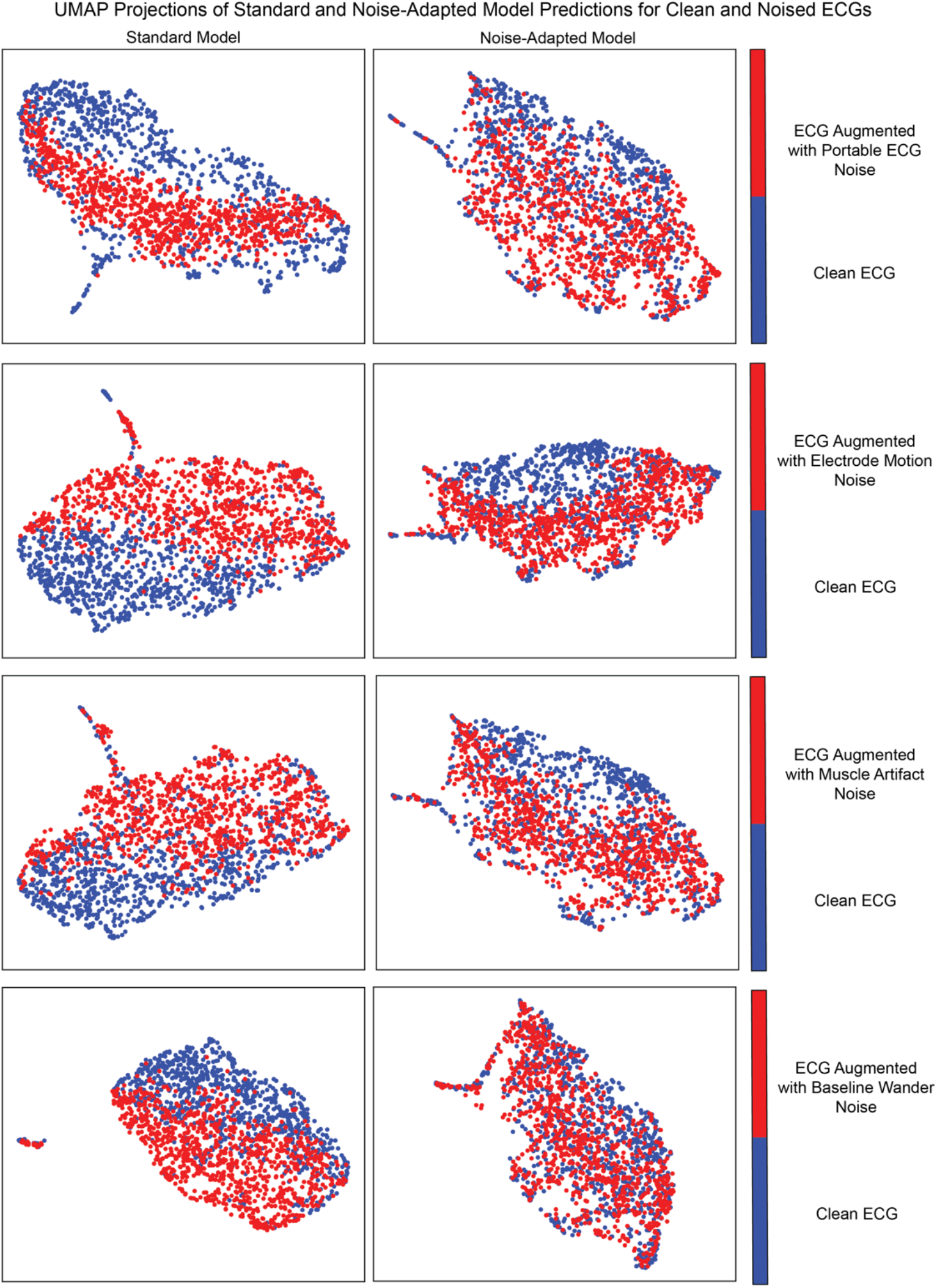
UMAP Projections of Last-Layer Predictions of Standard and Noise-Adapted Model for Clean and Noise-Augmented ECGs. Abbreviations: UMAP, uniform manifold approximation and projection; ECG, electrocardiogram.

Quantitatively, the scaled Euclidean distances between predictions for noised and clean versions of the ECGs were lower for the noise-adapted model across all four noises. In the ECG with noise derived from portable ECGs, for instance, the average scaled Euclidean distance for the standard model was 0.50 (95% CI 0.49-0.51) and for the noise adapted model was 0.41 (95% CI 0.40-0.42). Similarly for the baseline wander it was 0.52 (0.51-0.53) and 0.36 (0.36-0.37), respectively (**Table 3**).

**Table 3.**
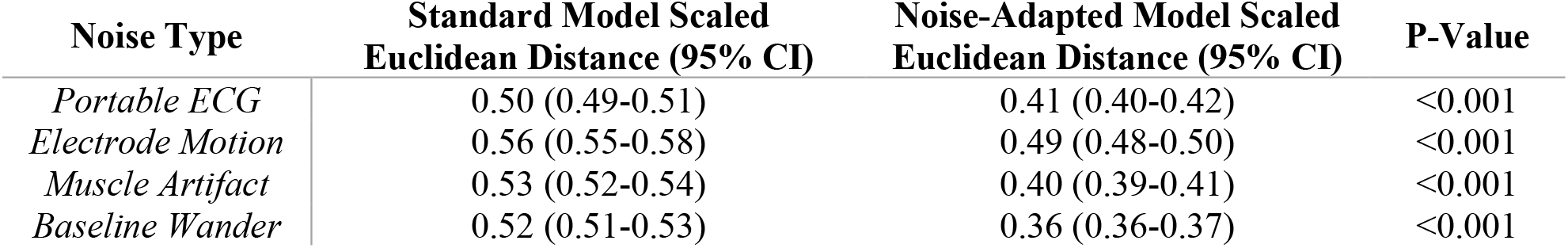
Scaled Euclidean distance between last-layer predictions of noise-adapted and standard models for noise-augmented and clean ECGs across different noise types at an SNR of 0.5. Abbreviations: SNR, signal-to-noise ratio; ECG, electrocardiogram; CI, confidence interval; P-Value, probability value.

## DISCUSSION

We developed a novel strategy that automates the detection of hidden signatures of cardiovascular diseases, such as LVSD, adapted for noisy single-lead ECGs obtained on wearable and portable devices. Using this approach, we developed a noise-adapted deep-learning algorithm that accurately identifies LV systolic dysfunction from single-lead ECG data and was resilient to significant noisy artifacts, despite not encountering the specific noises in the model development process. Specifically, the algorithm demonstrates excellent discriminatory performance even on ECGs containing twice as much noise as signal, features that make it ideal for wearable device-based screening strategies. Notably, the algorithm was developed and validated in a diverse population and demonstrates consistent performance across age, sex, and race subgroups. The noise-adapted approach defines a novel paradigm on how to build robust, wearable-ready, single-lead ECG cardiovascular screening models from clinical ECG repositories, with significant potential to expand the screening of LV structural cardiac disorders to low-resource settings with limited access to hospital-grade equipment.

Noise-adapted training of deep learning algorithms represents a relatively novel field of AI research, focused on expanding the use of AI tools to everyday life by accounting for noise and artifacts that may preclude their reliable use in this setting. Models trained on clinical ECGs have been applied to wearable device ECGs, but have traditionally shown significantly lower performance on wearable device ECGs than on held-out clinical ECG test sets.^6,7^ Due to the lack of publicly available wearable device ECG datasets, however, development of models for direct use with wearable ECG data remains infeasible. The current 12-lead ECG-based models are limited to investments by health systems to incorporate tools into digital ECG repositories, and thereby limited to individuals who already seek care in those systems. In addition to the clinically indicated ECGs limiting the scope of screening, even this technology may not be available or cost-effective for smaller hospitals and clinics with limited access to digital ECGs.

Wearable devices allow obtaining ECGs that are more accessible and allow for community-wide screening, an important next step in the early detection of common and rare cardiomyopathies. On this note, our approach represents a major advancement from a methodological and clinical standpoint. First, it augments clinical ECG datasets in such way that it enables reliable modelling of noisy, wearable-derived, single-lead ECG signals. Second, it demonstrates that through noise-augmentation, single-lead ECG models can retain the prognostic performance of 12-lead ECG models, as shown here for the task of predicting LV systolic dysfunction.

Our approach also offers a strategy to examine the process by which the models achieve better performance. Compared with the current approach of developing models, our noise-adapted approach resulted in selective removal of noise from signal, even for noises the model hadn’t encountered before, while preserving the model’s robustness in discerning complex hidden labels. This strategy is particularly important for a model intended for use on wearable devices, which capture ECGs in unpredictable settings with varying types and magnitudes of noise.

This study has several limitations. First, this model was developed using ECGs from patients who had both an ECG and a clinically indicated echocardiogram. Though this population differs from the intended broader real-world use of this algorithm as a screening method for LV systolic dysfunction among individuals with no clinical disease, the consistent performance across demographic subgroups suggests robustness and generalizability of the model’s performance. Nevertheless, prospective assessments in the intended screening setting are warranted. Second, the model performance may vary by the severity of LV systolic dysfunction. Though the LVEF threshold of 40% was selected due to its therapeutic implications,^14^ it is possible that the model performance among patients with an LVEF near to this cutoff differed compared with those individuals with LVEF significantly higher or lower than 40%. This might also be attributable to a lack of precision in LVEF measurement by echocardiography, which has shown to be less precise relative to other approaches, such as magnetic resonance imaging.^17,18^ Finally, four distinct types of randomly generated noise were used during training and randomly selected sequences of four real-world noises were used at multiple signal-to-noise ratios in the evaluation of performance on the test set. Though this suggests that the model performance generalizes well to unseen noise, we cannot ascertain whether it maintains performance on every type and magnitude of noise possible on wearable devices.

## CONCLUSIONS

We developed a novel strategy that automates the detection of hidden signatures of cardiovascular diseases, such as LVSD, adapted for noisy single-lead ECGs obtained on wearable and portable devices. Using this approach, we developed a single-lead ECG algorithm that accurately identifies LV systolic dysfunction despite significant noisy artifacts, suggesting a novel approach for the development of wearable adapted tools from clinical ECG repositories.

## METHODS

The study was reviewed by the Yale Institutional Review Board, which approved the study protocol and waived the need for informed consent as the study represents secondary analysis of existing data. The data cannot be shared publicly.

### Study design

The study was designed as a retrospective analysis of a cohort of 116,210 patients, 18 years of age or older, who underwent clinically indicated ECG with paired echocardiograms within 15 days of the index ECG at the Yale-New Haven Health hospital. To ensure the generalizability of our models, we applied no exclusion criteria, including patients of all sexes, races, and ethnicities (Table S1).

### Data Source and Population

Raw voltage data for lead I was isolated from 12-lead ECGs collected at the Yale-New Haven Hospital (YNHH) between 2015 and 2021. Lead I was chosen as it represents the standard lead obtained from wearable devices.^4^ Each clinical ECG was recorded as a standard 10-second, 12-lead recording with a sampling frequency of 500 Hz. These ECGs were predominantly recorded using Philips PageWriter and GE MAC machines. Patient identifiers were used to link ECGs with an accompanying transthoracic echocardiogram within 15 days of the ECG. These echocardiograms had been evaluated by expert cardiologists, and the LVEF defined in their interpretation was identified. If multiple echocardiograms were performed within the 15-day window, the one nearest to each ECG was used to define the LVEF for the model development and evaluation.

### Data Preprocessing

A standard preprocessing strategy was used to isolate signal from lead I of 12-lead ECGs, that included median pass filtering and scaling to millivolts. The Lead I signal was then isolated from each ECG, and a one second median filter was calculated for and subtracted from each single-lead ECG to remove baseline drift. The amplitudes of each sample in each recording were then divided by a factor of 1000 to scale the voltage recordings to millivolts.

### Isolation of Frequency-Banded Gaussian Noise

Random gaussian noise within four distinct frequency ranges was isolated to train the noise-adapted model. High pass and low pass filters were applied to five minutes of random gaussian noise to isolate noise within each of the frequency ranges, which included 3-12 Hz, 12-50 Hz, 50-100 Hz, and 100-150 Hz. Each frequency range was specifically selected to model an element of real-world ECG noises. 3-12 Hz models the motion artifact noises attributable to tremors, which occur within this frequency range.^12,13^ The 50-100 Hz domain reflects consistent electrode contact noise,^13^ while the 12-50 Hz and 100-150 Hz ranges contain the lower and higher frequency muscle noises, respectively.^9,13^ Additionally, the 50-100 and 100-150 Hz ranges each contain multiples of 50 and 60 Hz, the two mains frequencies used in ECG acquisition,^9^ which make up powerline interference noise.^9,13^ (**Figure 3**)

**Figure 3.**
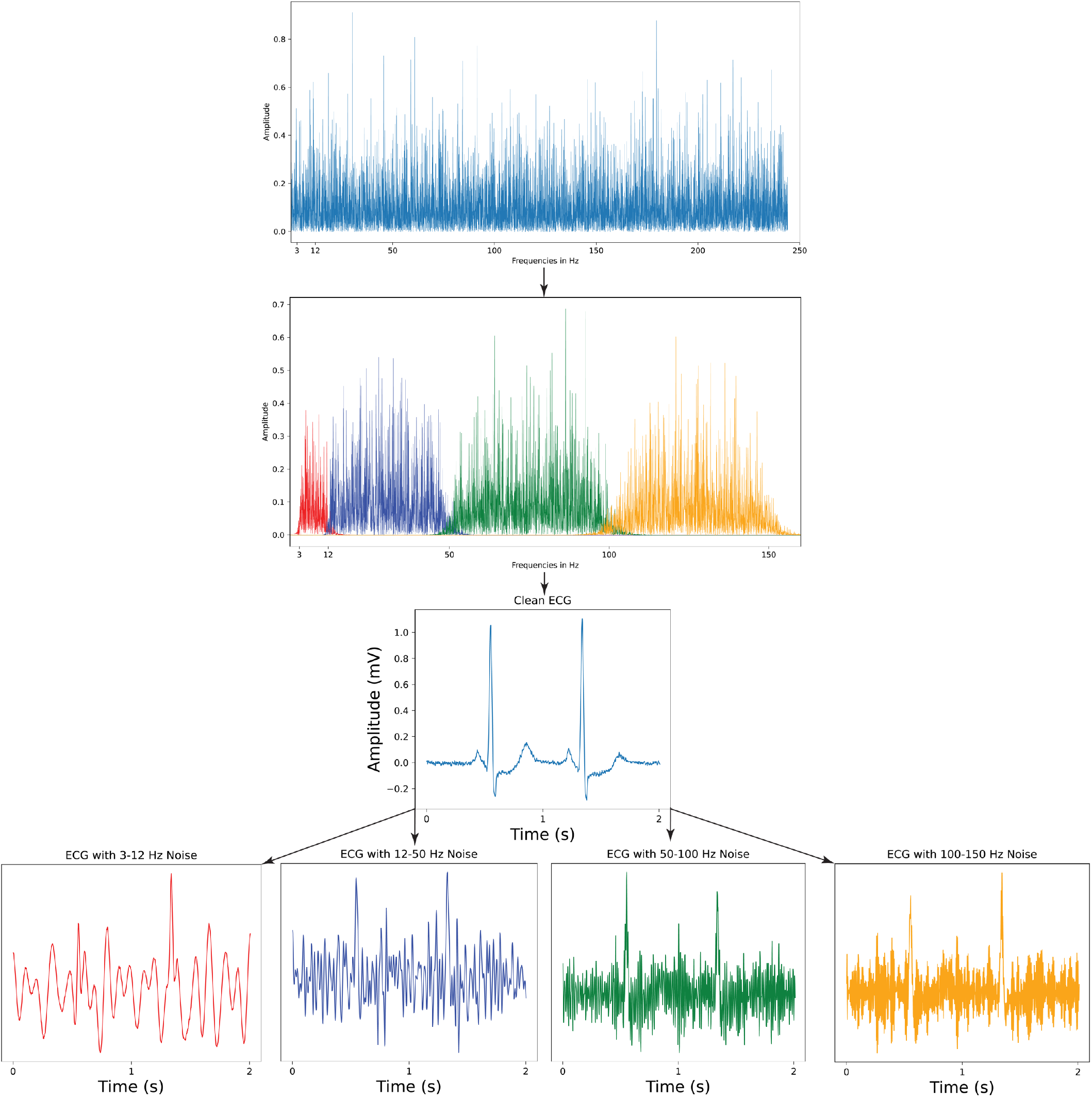
Noise Extraction and ECG Augmentation for Noise-Adapted Training. Abbreviations: ECG, electrocardiogram; Hz, hertz; s, seconds.

### Acquisition of Real-World, Public ECG Noise Recordings

Four real-world noise records, not included during training of either model, were used to test the models. These included three half-hour noise recordings from the MIT-BIH Noise Stress Test Database. The MIT-BIH dataset noises, each obtained at a sampling frequency of 360 Hz, represent three types of noises frequently encountered in ECGs: baseline wander noise, a low-frequency noise produced by lead or subject movement,^9^ muscle artifact, caused by muscle contractions,^13^ and electrode motion artifact, which is caused by irregular movement of the electrodes during ECG recordings.^9,13^ Each of these recordings were obtained using a standard 12-lead ECG recorder by positioning the electrodes on patient limbs such that the patients’ ECG signals were not visible in the recordings (**Figure 4**).^9^

**Figure 4.**
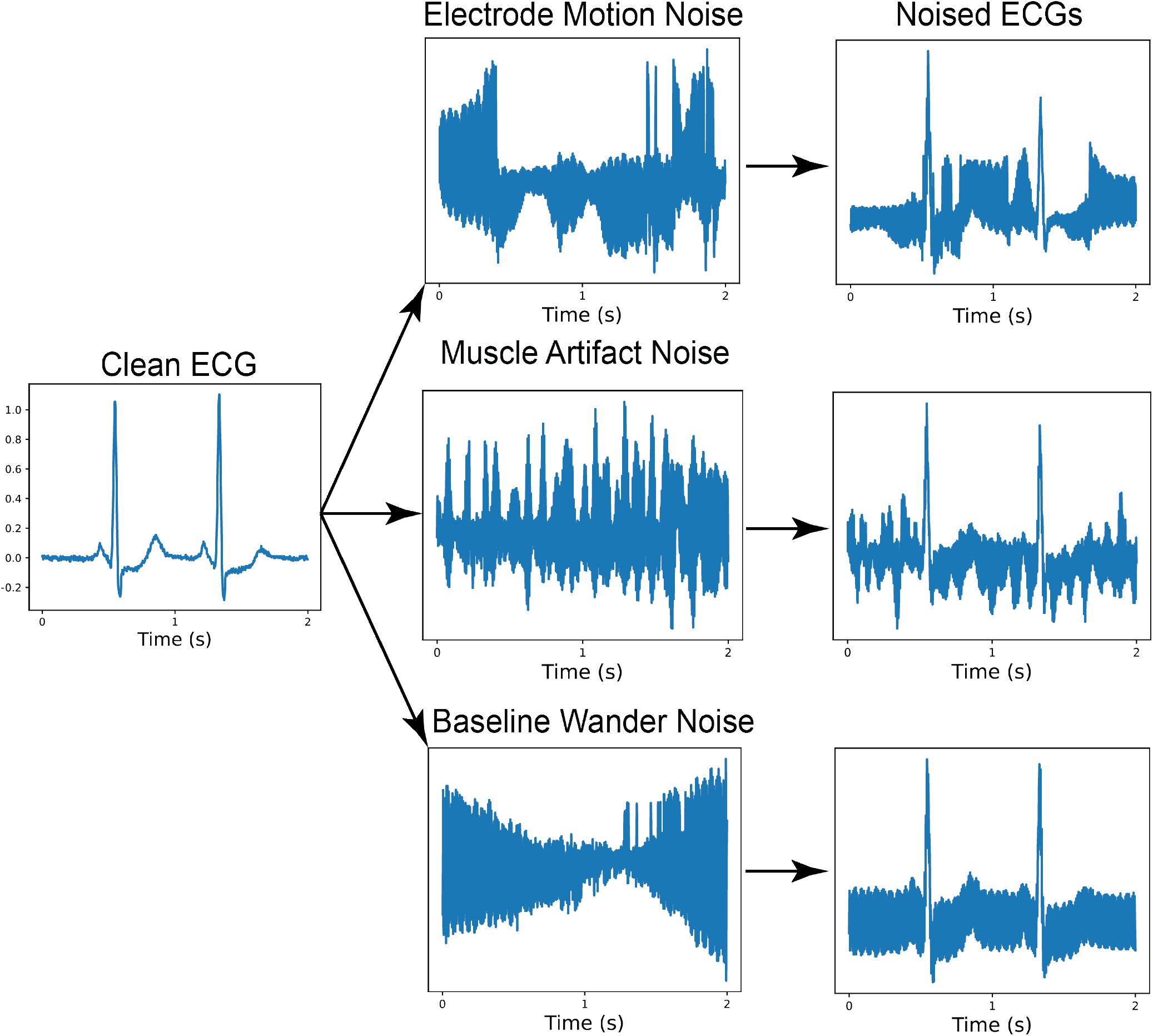
Noise Augmentation of Test Set with MIT-BIH Real-World Noises. Abbreviations: ECG, electrocardiogram; s, seconds.

### Noise Extraction from a Portable Device ECG

Real-world noise was also isolated from a 30-second, 300 Hz portable device ECG recording obtained using a KardiaMobile 6L portable ECG device. The noise was extracted from the recording by applying a modified version of the Fourier transform-based approach previously used to denoise ECGs.^19^ First, a fast Fourier transform (FFT) was applied to a 30-second ECG recording and the result was plotted in the frequency domain. Then, a threshold was manually selected to separate the high- and low-amplitude frequencies which contained signal and noise, respectively. Finally, instead of computing the inverse FFT on the frequencies with amplitudes greater than this threshold, an inverse FFT was applied to all frequencies with amplitudes below the selected threshold, yielding the noise (**Figure 5**).

**Figure 5.**
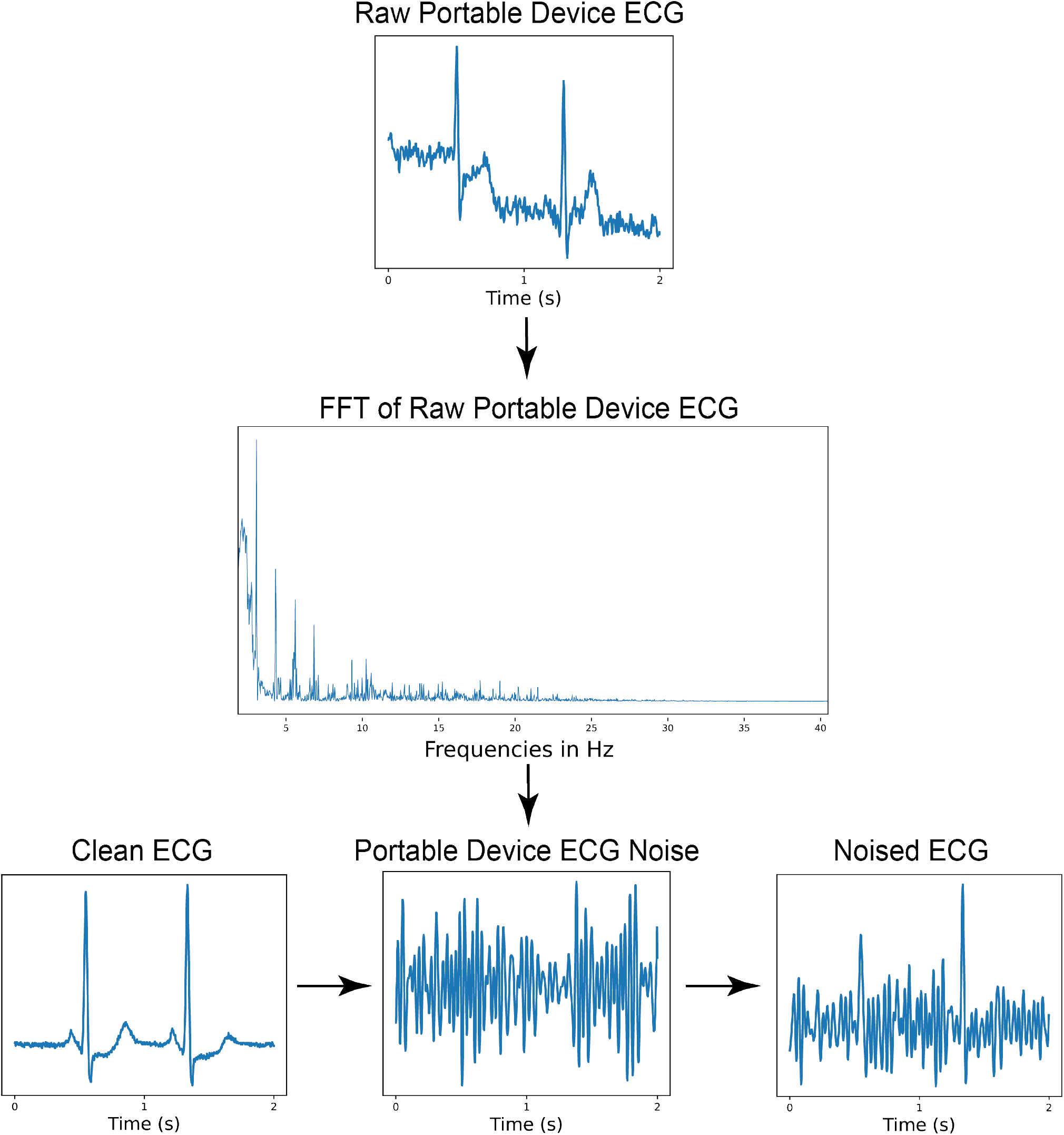
Portable ECG Device Noise Isolation and ECG Noise Augmentation. Abbreviations: ECG, electrocardiogram; s, seconds; FFT, Fast Fourier Transform; Hz, hertz.

### Noise Augmentation

While training the noise-adapted model, each ECG was included in the training dataset twice, with one of the four frequency ranges for the generated noise randomly selected each time an ECG was loaded. A 10-second sequence was then randomly selected from the full five-minute length of the selected noise recording. This noise sequence was then added to the clean ECG at a signal-to-noise ratio (SNR) randomly selected from a set of SNRs, including 0.50, 0.75, 1.00, and 1.25.^20^

During model evaluation, the noised versions of the test set were generated by following the same procedure as with the noised training set with several key modifications. First, the noise added to ECGs in the test set was sourced from either the baseline wander, electrode motion, or muscle artifacts noise recordings from the MIT-BIH dataset or from the 30-second noise sample from the KardiaMobile 6L portable ECG device. Second, the MIT-BIH noises and the portable ECG noise were all up-sampled from their original sampling frequencies, 360 Hz and 300 Hz, respectively, to a 500 Hz sampling frequency to match that of the clinical device ECGs. Third, the specific 10-second sequence of noise added to each ECG in the test set was randomly selected once and defined for each ECG, ensuring that every time any model was tested at any SNR for any specific noise, each individual ECG was always loaded with the same randomly selected sequence of noise. Model performance was evaluated separately for a larger set of SNRs, including all the SNRs used in training and SNRs of 1.50, 1.75, and 2.00.

### Outcome label

Each ECG included in the dataset had a corresponding LVEF value from a paired echocardiogram within 15 days of the ECG. The cutoff for low LVEF was set as LVEF < 40%, a threshold present in most heart failure diagnosis guidelines,^14^ and consistent with prior work in this space.^1,15^

### Model Training

All unique patients represented in the set of ECGs were then randomly subset on the patient level into training, validation, and held-out test sets (85%, 5%, 10%). We built and tested multiple convolutional neural network (CNN) models with varying numbers and sizes of convolutional layers and total model parameters. We selected the architecture yielding the highest area under the receiver operator characteristic (AUROC) curve on the validation set with the fewest number of parameters for the standard model. This architecture consisted of a (5000, 1, 1) input layer, corresponding to a 10-second, 500 Hz, Lead I ECG, followed by seven two-dimensional convolutional layers, each of which were followed by a batch normalization layer,^21^ ReLU activation layer, and a two-dimensional max-pooling layer. The output of the seventh convolutional layer was then taken as input into a fully connected network consisting of two dense layers, each of which were followed by a batch normalization layer, ReLU activation layer, and a dropout layer with a dropout rate of 0.5.^22^ The output layer was a dense layer with one class and a sigmoid activation function. Model weights were calculated for the loss function such that learning was not affected by the lower frequency of LVEF < 40% compared to the incidence of LVEF ≥ 40% using the effective number of samples class re-weighting scheme.^23^ (**Figure S2)**.

Both models were trained on the Keras framework in TensorFlow 2.9.1 and Python 3.9 using the Adam optimizer. First, the models were trained at a learning rate of 0.001 for one epoch. The learning rate was then lowered to 0.0001 and training was continued until performance on the validation set did not improve for three consecutive epochs. The epoch with the highest performance on the validation set was selected for each model.

### Learning Representation Assessments for Noise-Adapted Model

To visualize the variation between predictions for clean and noise-augmented data for each model, we first modified both the standard and noise-adapted models by removing their final output layers so both models instead produced the 320-dimensional vector output of the model’s final fully connected layer. We then randomly selected a 1,000 ECG subset from the held-out test set. For each of the two models, we generated the 320-dimension prediction vectors five times for each of the 1,000 ECGs—once without noise, and once augmented at an SNR of 0.5 for each of the four noises used for testing. We then visualized the variation in the predictions separately for the standard and noise-adapted models using uniform manifold approximation and projection (UMAP), which constructs a two-dimensional representation of the 320-dimension prediction vectors.^16^ The variation in predictions was numerically assessed by the pair-wise calculation of the Euclidean distances between the 320-dimensional prediction vectors for the clean and noised data for each of the four noises. These Euclidean distances were then scaled on a per-model and per-noise basis by dividing by the total range of pair-wise Euclidean distances for each model and noise combination. The average scaled Euclidean distance and a 95% confidence interval was then calculated for each model and noise combination.

### Statistical Analysis

Summary statistics are presented as counts (percentages) and median (interquartile range, IQR), for categorical and continuous variables, respectively. Model performance was evaluated in the held-out test set both with and without added real-world noises. We used area under receiving operation characteristics (AUROC) to measure model discrimination. We also assessed area under precision recall curve (AUPRC), sensitivity, specificity, positive predictive value (PPV), negative predictive value (NPV), and diagnostic odds ratio, and chose threshold values based on cutoffs that achieved sensitivity of 0.90 in validation data. We used a DeLong’s test to compare AUROCs of the noise-adapted and standard models for each noise at each signal-to-noise ratio.^24^ 95% confidence intervals for AUROC were calculated using DeLong’s algorithm.^24,25^ A paired t-test was used to compute the probability of overlap for the scaled Euclidean distances between last-layer projections of the noise-adapted and standard models. All analyses were performed using Python 3.9 and level of significance was set at an alpha of 0.05.

## Data Availability

All data produced in the present work are contained in the manuscript

## Author contributions

RK conceived the study and accessed the data. AK and RK developed the model. AK, VS, and RK pursued the statistical analysis. AK and EKO drafted the manuscript. All authors provided feedback regarding the study design and made critical contributions to writing of the manuscript. RK supervised the study, procured funding, and is the guarantor.

