## Supplement for "Detection of Left Ventricular Systolic Dysfunction from Single-Lead Electrocardiography Adapted for Wearable Devices"

**Table S1. Patient-level baseline characteristics of study population.** Data presented median [IQR] for age and number (percent) for other variables.

| **Characteristic** | **Total (N = 116,210)** |
| --- | --- |
| **Sex** |  |
| Female | 50,776 (43.7%) |
| Male | 53,271 (45.8%) |
| Unknown | 12,149 (10.5%) |
| **Age (years)** | 68 [56-78] |
| **Race** |  |
| Hispanic | 9,349 (8.0%) |
| White | 75,928 (65.3%) |
| Black | 14,000 (12.0%) |
| Other | 16,843 (14.5%) |

**Table S2. Performance of noise-adapted and standard model on different noise types at each signal-to-noise ratio on the held-out test set.** Abbreviations: PPV, positive predictive value; NPV, negative predictive value; AUROC, area under receiver operating characteristic curve; AUPRC, area under precision recall curve; OR, odds ratio.

| **Model** | **Noise** | **SNR** | **PPV** | **NPV** | **Specificity** | **Sensitivity** | **AUROC (95% CI)** | **AUPRC** |
| --- | --- | --- | --- | --- | --- | --- | --- | --- |
| Noise-Adapted | Clean | N/A | 0.203 | 0.990 | 0.676 | 0.924 | 0.896 (0.886-0.905) | 0.455 |
|  | Portable ECG | 0.50 | 0.152 | 0.993 | 0.523 | 0.956 | 0.871 (0.861-0.882) | 0.392 |
|  | Portable ECG | 0.75 | 0.165 | 0.992 | 0.569 | 0.951 | 0.879 (0.868-0.889) | 0.411 |
|  | Portable ECG | 1.00 | 0.172 | 0.992 | 0.592 | 0.946 | 0.883 (0.872-0.893) | 0.421 |
|  | Portable ECG | 1.25 | 0.177 | 0.992 | 0.607 | 0.945 | 0.885 (0.875-0.895) | 0.428 |
|  | Portable ECG | 1.50 | 0.181 | 0.992 | 0.619 | 0.942 | 0.887 (0.877-0.897) | 0.433 |
|  | Portable ECG | 1.75 | 0.185 | 0.992 | 0.628 | 0.942 | 0.888 (0.878-0.898) | 0.436 |
|  | Portable ECG | 2.00 | 0.188 | 0.992 | 0.636 | 0.940 | 0.889 (0.880-0.899) | 0.439 |
|  | Electrode Motion | 0.50 | 0.126 | 0.993 | 0.395 | 0.971 | 0.858 (0.846-0.869) | 0.361 |
|  | Electrode Motion | 0.75 | 0.138 | 0.994 | 0.459 | 0.968 | 0.870 (0.859-0.881) | 0.388 |
|  | Electrode Motion | 1.00 | 0.147 | 0.993 | 0.501 | 0.961 | 0.876 (0.865-0.886) | 0.402 |
|  | Electrode Motion | 1.25 | 0.154 | 0.992 | 0.531 | 0.955 | 0.879 (0.869-0.889) | 0.410 |
|  | Electrode Motion | 1.50 | 0.160 | 0.992 | 0.553 | 0.953 | 0.881 (0.871-0.892) | 0.416 |
|  | Electrode Motion | 1.75 | 0.165 | 0.992 | 0.569 | 0.950 | 0.883 (0.873-0.893) | 0.422 |
|  | Electrode Motion | 2.00 | 0.167 | 0.991 | 0.579 | 0.945 | 0.885 (0.875-0.895) | 0.426 |
|  | Muscle Artifact | 0.50 | 0.175 | 0.989 | 0.608 | 0.927 | 0.871 (0.861-0.882) | 0.389 |
|  | Muscle Artifact | 0.75 | 0.182 | 0.990 | 0.626 | 0.930 | 0.879 (0.869-0.890) | 0.415 |
|  | Muscle Artifact | 1.00 | 0.186 | 0.990 | 0.636 | 0.931 | 0.884 (0.873-0.894) | 0.429 |
|  | Muscle Artifact | 1.25 | 0.190 | 0.991 | 0.643 | 0.934 | 0.886 (0.877-0.896) | 0.437 |
|  | Muscle Artifact | 1.50 | 0.192 | 0.991 | 0.649 | 0.932 | 0.888 (0.879-0.898) | 0.444 |
|  | Muscle Artifact | 1.75 | 0.194 | 0.991 | 0.652 | 0.934 | 0.890 (0.880-0.899) | 0.448 |
|  | Muscle Artifact | 2.00 | 0.194 | 0.990 | 0.654 | 0.930 | 0.891 (0.881-0.900) | 0.449 |
|  | Baseline Wander | 0.50 | 0.186 | 0.990 | 0.634 | 0.932 | 0.883 (0.873-0.893) | 0.427 |
|  | Baseline Wander | 0.75 | 0.192 | 0.990 | 0.651 | 0.926 | 0.887 (0.877-0.897) | 0.440 |
|  | Baseline Wander | 1.00 | 0.195 | 0.990 | 0.658 | 0.926 | 0.889 (0.879-0.899) | 0.448 |
|  | Baseline Wander | 1.25 | 0.198 | 0.990 | 0.664 | 0.926 | 0.890 (0.881-0.900) | 0.453 |
|  | Baseline Wander | 1.50 | 0.199 | 0.990 | 0.666 | 0.928 | 0.891 (0.881-0.901) | 0.455 |
|  | Baseline Wander | 1.75 | 0.200 | 0.991 | 0.668 | 0.929 | 0.892 (0.882-0.902) | 0.457 |
|  | Baseline Wander | 2.00 | 0.201 | 0.991 | 0.669 | 0.929 | 0.892 (0.883-0.902) | 0.457 |
| Standard | Clean | N/A | 0.207 | 0.988 | 0.688 | 0.910 | 0.895 (0.884-0.905) | 0.475 |
|  | Portable ECG | 0.50 | 0.091 | 0.981 | 0.132 | 0.972 | 0.723 (0.706-0.739) | 0.200 |
|  | Portable ECG | 0.75 | 0.096 | 0.987 | 0.182 | 0.973 | 0.763 (0.748-0.779) | 0.242 |
|  | Portable ECG | 1.00 | 0.102 | 0.990 | 0.233 | 0.974 | 0.788 (0.773-0.802) | 0.270 |
|  | Portable ECG | 1.25 | 0.108 | 0.990 | 0.287 | 0.969 | 0.805 (0.791-0.819) | 0.291 |
|  | Portable ECG | 1.50 | 0.114 | 0.990 | 0.331 | 0.964 | 0.818 (0.805-0.831) | 0.306 |
|  | Portable ECG | 1.75 | 0.120 | 0.991 | 0.369 | 0.961 | 0.827 (0.814-0.840) | 0.318 |
|  | Portable ECG | 2.00 | 0.125 | 0.991 | 0.398 | 0.958 | 0.834 (0.822-0.847) | 0.329 |
|  | Electrode Motion | 0.50 | 0.088 | 0.988 | 0.079 | 0.990 | 0.792 (0.779-0.806) | 0.239 |
|  | Electrode Motion | 0.75 | 0.101 | 0.991 | 0.219 | 0.978 | 0.815 (0.802-0.829) | 0.264 |
|  | Electrode Motion | 1.00 | 0.114 | 0.991 | 0.330 | 0.965 | 0.829 (0.817-0.842) | 0.285 |
|  | Electrode Motion | 1.25 | 0.127 | 0.990 | 0.414 | 0.954 | 0.839 (0.827-0.851) | 0.301 |
|  | Electrode Motion | 1.50 | 0.137 | 0.989 | 0.468 | 0.942 | 0.846 (0.834-0.858) | 0.313 |
|  | Electrode Motion | 1.75 | 0.145 | 0.989 | 0.508 | 0.934 | 0.851 (0.839-0.862) | 0.324 |
|  | Electrode Motion | 2.00 | 0.152 | 0.988 | 0.537 | 0.925 | 0.855 (0.843-0.866) | 0.333 |
|  | Muscle Artifact | 0.50 | 0.100 | 0.991 | 0.210 | 0.979 | 0.807 (0.795-0.820) | 0.245 |
|  | Muscle Artifact | 0.75 | 0.124 | 0.993 | 0.385 | 0.970 | 0.832 (0.820-0.844) | 0.274 |
|  | Muscle Artifact | 1.00 | 0.143 | 0.991 | 0.488 | 0.951 | 0.844 (0.833-0.855) | 0.294 |
|  | Muscle Artifact | 1.25 | 0.157 | 0.990 | 0.546 | 0.940 | 0.851 (0.840-0.863) | 0.310 |
|  | Muscle Artifact | 1.50 | 0.166 | 0.989 | 0.584 | 0.929 | 0.857 (0.846-0.868) | 0.323 |
|  | Muscle Artifact | 1.75 | 0.175 | 0.989 | 0.612 | 0.923 | 0.861 (0.850-0.872) | 0.335 |
|  | Muscle Artifact | 2.00 | 0.181 | 0.988 | 0.631 | 0.912 | 0.864 (0.853-0.875) | 0.344 |
|  | Baseline Wander | 0.50 | 0.095 | 0.993 | 0.158 | 0.987 | 0.802 (0.789-0.816) | 0.236 |
|  | Baseline Wander | 0.75 | 0.113 | 0.990 | 0.324 | 0.964 | 0.822 (0.810-0.835) | 0.260 |
|  | Baseline Wander | 1.00 | 0.130 | 0.989 | 0.433 | 0.946 | 0.833 (0.821-0.846) | 0.277 |
|  | Baseline Wander | 1.25 | 0.144 | 0.988 | 0.503 | 0.933 | 0.841 (0.829-0.853) | 0.292 |
|  | Baseline Wander | 1.50 | 0.154 | 0.987 | 0.546 | 0.923 | 0.847 (0.835-0.859) | 0.303 |
|  | Baseline Wander | 1.75 | 0.163 | 0.988 | 0.578 | 0.918 | 0.851 (0.840-0.863) | 0.314 |
|  | Baseline Wander | 2.00 | 0.169 | 0.986 | 0.599 | 0.908 | 0.855 (0.843-0.866) | 0.322 |

**Figure S1. Flow chart of study cohort and analysis.**

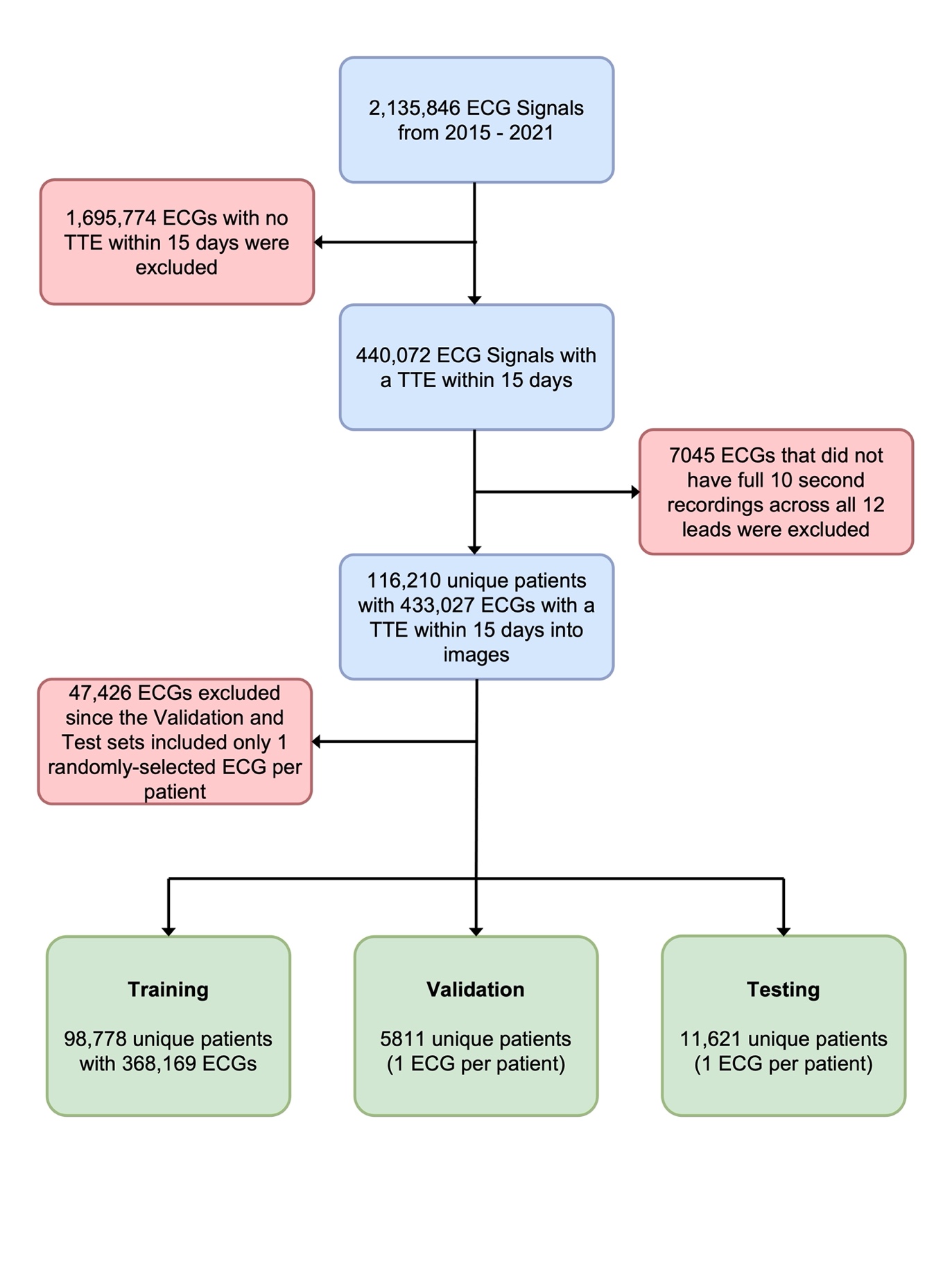

**Figure S2. Model architecture used for model development.**

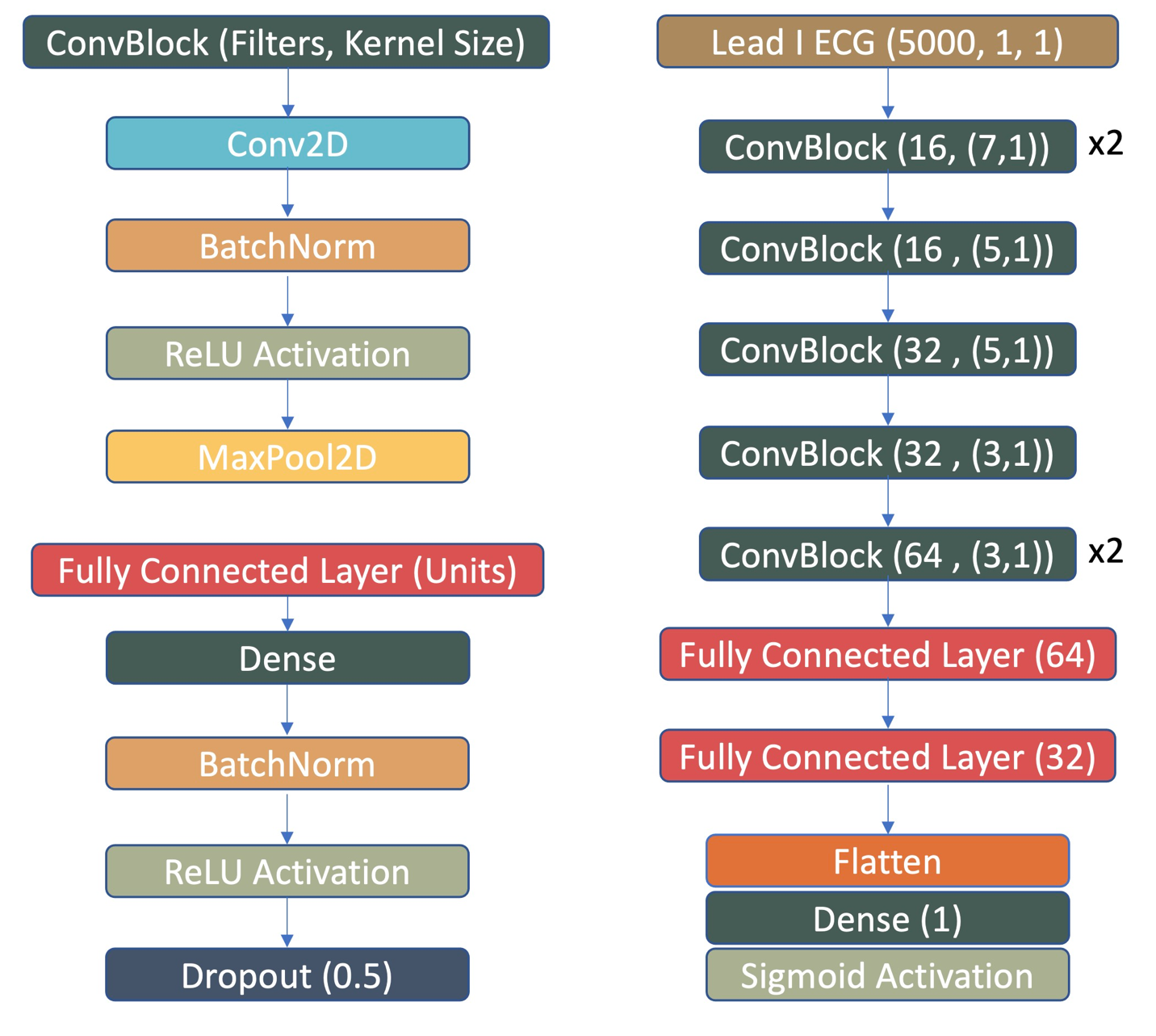
